# Whence the next pandemic? The intersecting global geography of the animal-human interface, poor health systems and air transit centrality reveals conduits for high-impact spillover

**DOI:** 10.1101/2020.07.27.20163196

**Authors:** Michael G. Walsh, Shailendra Sawleshwarkar, Shah Hossain, Siobhan M. Mor

## Abstract

The health and economic impacts of infectious disease pandemics are catastrophic as most recently manifested by coronavirus disease 2019 (COVID-19). The emerging infections that lead to substantive epidemics or pandemics are typically zoonoses that cross species boundaries at vulnerable points of animal-human interface. The sharing of space between wildlife and humans, and their domesticated animals, has dramatically increased in recent decades and is a key driver of pathogen spillover. Increasing animal-human interface has also occurred in concert with both increasing globalisation and failing health systems, resulting in a trifecta with dire implications for human and animal health. Nevertheless, to date we lack a geographical description of this trifecta that can be applied strategically to pandemic prevention. This investigation provides the first geographical quantification of the intersection of animal-human interfaces, poor human health system performance and global connectivity via the network of air travel. In so doing, this work provides a systematic, data-driven approach to classifying spillover hazard based on the distribution of animal-human interfaces while simultaneously identifying globally connected cities that are adjacent to these interfaces and which may facilitate global pathogen dissemination. We present this geography of high-impact spillover as a tool for developing targeted surveillance systems and improved health infrastructure in vulnerable areas that may present conduits for future pandemics.

## Introduction

The global spread of severe acute respiratory syndrome coronavirus-2 (SARS-CoV-2) in 2020 has shown how rapidly emerging infectious diseases can devastate human health and national economies. Such infections typically have zoonotic origins, with other notable examples being SARS-CoV in 2002^1^, influenza A H1N1 virus in 2009^2^, and the West African Ebola virus disease epidemic of 2013-2016^3^. Emerging infectious diseases are increasing in incidence and expanding in geographic range due in large part to direct and diffuse anthropogenic pressure across ecosystems^4^. This is particularly true in areas with high wildlife biodiversity that are experiencing land-use changes such as deforestation ^4,5,6,7,^ although the process of disease emergence is complex and cannot be attributed to any one driver.

Morse et al. have developed a useful framework for conceptualising a staged process of pathogen emergence in humans^8^. In the first stage, potential pathogens circulate exclusively in reservoir hosts. While this stage precedes spillover, the stage is still highly influenced by human activities that place stress on animal populations, such as habitat destruction, population displacement, and nutritional insecurity, thereby altering pathogen circulation in affected non-human host populations^9^. Various researchers have hypothesized that certain animal taxa, such as bats^8^, harbor a significantly higher proportion of zoonotic viruses. However, differential impact of specific taxa on spillover has been recently challenged by research showing that overall species richness (rather than taxonomy) is the primary driver of pathogen richness among mammalian and bird reservoir hosts^10^.

In the framework’s second stage, activities that increase animal-human contact (such as forest management practices or raising domestic animals) create opportunities for spillover from reservoir hosts into human populations. The risk of spillover appears greatest both from generalist species that are highly abundant and adapted to human-dominated landscapes, and – conversely – from those specialist species that are threatened specifically due to habitat loss and human exploitation^9^; both scenarios confirm the central influence of humans as a driving force behind spillover. Hassell et al. argued that wildlife–livestock–human interfaces emerging under urbanisation represent particularly critical points for cross-species transmission, noting an urgent need for better characterisation of peri-urban wildlife interfaces using transdisciplinary approaches^11^. Despite the critical influence of anthropogenic pressure, spillover is a complex process requiring that pathogens overcome many landscape and immunological barriers^12^, and even when successful, not all spillovers lead to sustained transmission. Most often this is due to a lack of efficient transmission in the new host populations. Occasionally, spillover into naïve human or domesticated animal populations does result in efficient transmission.

The framework’s final stage is marked by sustained onward transmission and widespread regional or global dissemination. At this stage, the capacity of local health systems to rapidly detect cases of a novel disease and control ongoing chains of transmission is key to preventing broader dissemination from the original focus^8^. In this work we distinguish wildlife-origin zoonoses with the potential to transmit efficiently between humans as *impactful spillovers*, and we further designate impactful spillovers with the potential to disseminate rapidly to regions beyond their origin focus as *high-impact spillovers*. High-impact spillovers are therefore a function of the capacity of local health systems and the proximity of the initial spillover to conduits of broader global dissemination, particularly transportation hubs such as airports^8^. Consequently, in order to block emerging zoonoses with pandemic potential (high-impact spillovers), biosurveillance systems must simultaneously consider critical animal-human interfaces, the performance and reach of the health systems, and the biosecurity of proximate transportation hubs that can serve as conduits for rapid global dissemination.

Despite the documented importance of animal-human interfaces for zoonotic transmission of pathogens, and the framework described above for understanding the staged transition from spillover through to human pandemic, we still lack a practical, data-driven and synthetic description of the geography of the critical intersection of animal-human interface and vulnerable points with low disease detection and rapid widespread dissemination potential. The aims of the current work were therefore to (1) describe and quantify the global geography of the interfaces between mammalian and bird wildlife and humans and their domestic livestock; and (2) to synthesize the geography of the wildlife-livestock/poultry-human interface, poor health system performance, and the global network of air travel to identify cities whose global connectedness and proximity to animal-human interfaces indicate significant potential to serve as conduits for high-impact spillover.

## Methods

### Data sources

Raster data for mammalian and bird species richness, livestock and poultry densities, and human population density were acquired to describe the intersection of their geographic distributions as landscapes of potential animal-human interface. Mammalian and bird species richness were used as a representation of total mammalian and bird diversity (⍰-diversity), respectively, rather than distinguishing between taxonomic groups since it has recently been shown that overall species richness is the primary driver of pathogen richness among hosts rather than taxonomy^10^. A mammalian richness raster was acquired from the Socioeconomic Data and Applications Center (SEDAC) repository^13^. Species richness was quantified using the geographic extents of the International Union for Conservation of Nature (IUCN) assessment of mammalian species (5,488 species in 156 families). A total bird richness raster was obtained from the biodiversity mapping project^14^ and was constructed from the Handbook of Birds of the World^15^, BirdLife International^16^, and IUCN. The Gridded Livestock of the World (GLW) provided livestock densities for cattle, pigs, sheep, goats, and buffaloes, and poultry densities for chicken and ducks^17^. The GLW was updated in 2018 to more appropriately account for spatial heterogeneity within and between countries. Aggregate livestock and poultry rasters were created by taking the sum of the absolute number of cattle, pigs, sheep, goats, and buffaloes per unit area, and chicken and ducks per unit area, respectively, since the current aim was to describe the geography of the wild mammal-livestock-human interface, and wild bird-poultry-human interfaces. Human population density was also obtained from the SEDAC repository and is derived from the Global Rural-Urban Mapping Project version 4 estimates for the 2015 population^18^. Finally, defining high-impact spillover potential requires a measure of the distribution of health system performance as an indication of the local capacity to detect and control the occurrence of cases of a novel zoonosis. The infant mortality ratio (IMR) was chosen as a proxy for health system performance because this has been recognised as a robust and widely used indicator of health infrastructure and health system performance and used extensively to compare health services between countries^19,20^. The IMR has been shown to correlate very well with disability adjusted life expectancy (DALE) as well as the Human Development Index (HDI) and the Inequality-Adjusted Human Development Index (IHDI) and strongly relates to structural issues that affect entire populations, such as economic development, general living conditions, social well-being, and the quality of the environment^19,21^. The raster of the IMR was obtained from SEDAC^22^. Mammalian and bird richness, human population, and IMR rasters were aggregated to the spatial resolution of the livestock and poultry rasters such that all features were analysed with a granularity of 5 arc minutes, which is approximately 10 km. Finally, an adjacency matrix for the global airport network was acquired from WorldPop to compute the network centrality of individual airports within the global network^23^. This data product was constructed by WorldPop to model annual air passenger flows and comprises 1491 airports (network nodes) with 644,406 flight connections between them (network edges)^24^.

### Data analysis

The global distribution of each landscape feature is presented in the left column of S1 Figure 1, with the corresponding top quartile of each distribution (75th percentile) presented in the right column. The objective was to demarcate geographic zones based on the degree of intersection of each landscape feature, thus constructing proxy interfaces between humans, livestock, poultry, and mammalian and bird wildlife, and finally identifying where these interfaces intersect with poor health system performance. Wild bird-poultry and wild mammal-livestock interfaces were considered separately as there is evidence that interfaces arise largely from interaction between phylogenetically related and/or sympatric species^25^. Intersection was classified as the co-occurrence of the top quartile (75th percentile) of each specified feature. The classification scheme was also re-constructed using the co-occurrence of the 50^th^ percentile for each landscape feature to examine how different distribution classifications affect the demarcation of interface and associated hazard.

**Figure 1.**
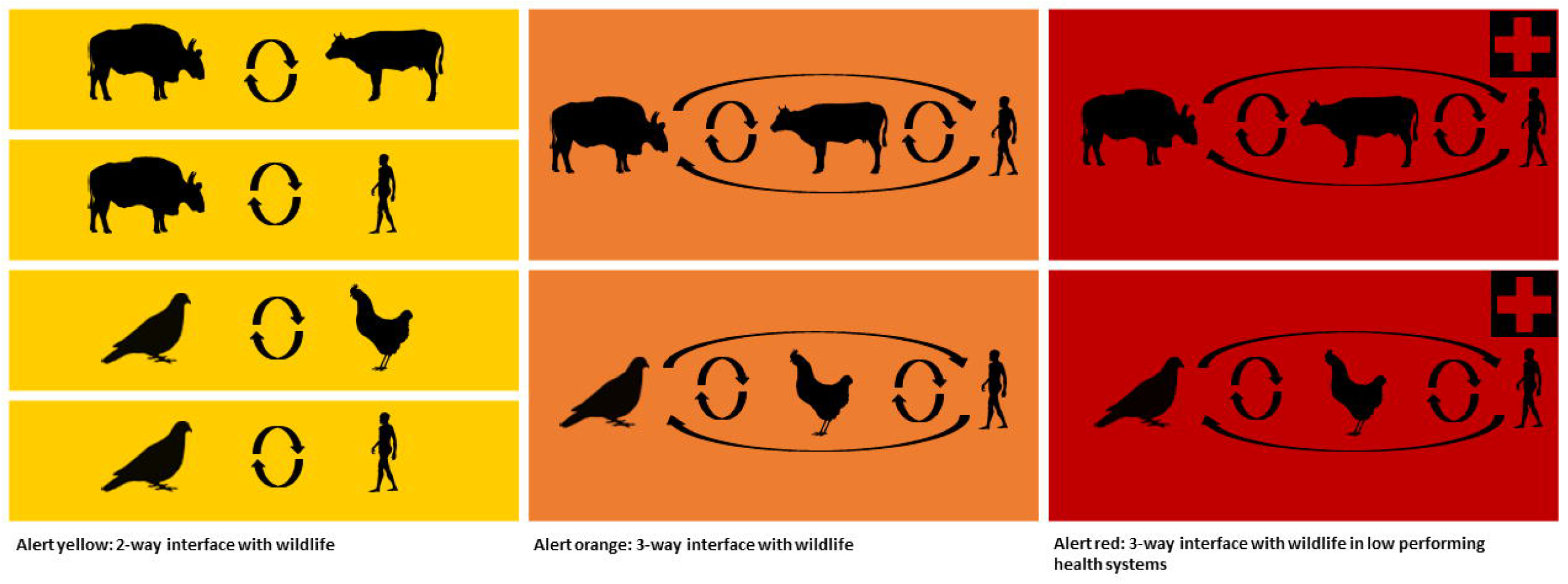
Alert-level hierarchy of the animal-human interface. All three zones represent landscapes of high anthropogenic pressure intersecting with high biodiversity, and thus all three zones indicate potential for impactful spillover. The transition from yellow to orange marks increasing pressure on, and contact with, wildlife populations and thus increasing risk of spillover. The transition from orange to red marks the intersection of high animal-human interface with poor health system performance and thus the areas of highest risk for high-impact spillover.

Three alert levels (yellow, orange, and red) were identified, as conceptualised in Figure 1. All levels represent landscapes with both high anthropogenic pressure and high biodiversity, and thus have potential for impactful spillover. Alert-level yellow depicts two-way interfaces between mammalian/bird richness and human population or livestock/poultry densities. Alert-level orange depicts three-way interfaces between mammalian/bird richness, human population density, and livestock/poultry densities. Alert-level red depicts the same animal-human interfaces as alert-level orange, but extends the intersection of these interfaces to include the top quartile of infant mortality. Therefore, the hierarchy of this classification system designates two-way interfaces with wildlife (yellow) as the minimum source requirement for high-impact spillovers, whereas alert-level orange represents landscapes with maximal pressure on, and contact with, wildlife populations. Finally, alert-level red depicts where the geography of maximum human-animal interface intersects with the geography of poor health system performance. The latter represents those areas that might be expected to miss detection of early cases of spillover to humans and thus presents the greatest risk for high-impact spillover. This scheme thus builds on the work by Morse et al. by 1) quantifying and mapping the degree of shared space between animals and humans, which they posit as driving the first 2 stages of their framework, and 2) quantifying and mapping the degree of health system capacity to intercept global dissemination of zoonotic pathogens that demonstrate onward human-to-human transmission, which defines the third stage of their framework.

The validity of these animal-human interface metrics were tested using two case studies. The value of these metrics lies in how well they represent contact between wildlife, livestock/poultry, and humans, and the extent to which they intersect with poor health system performance. Therefore, the two diseases selected as case studies were chosen based on their representation of infection ecology at the relevant animal-human interfaces and not based on their potential to cause pandemics in humans. As such anthrax was selected because it is a model disease for the mammal-livestock-human interface^26,27,^ while highly pathogenic avian influenza A H5N1 (HPAI H5N1) was selected as a similarly good representation of the bird-poultry-human interface^28^. The Food and Agriculture Organisation’s Global Animal Disease Information System (EMPRES-i) was used to capture reported occurrences of anthrax and HPAI H5N1 between 1 January, 2010, and 1 May 2020^29^. The EMPRES-i system has a particular focus on transboundary animal diseases and emergent zoonoses. Only those occurrences for which the exact location, or the location of the centroid of the subdistrict of the occurrence, was known were included in the case studies to minimise spatial uncertainty. For the two cases studies, respectively, the anthrax (n = 194) and HPAI H5N1 (n = 3247) cases were considered as point processes^30^. An inhomogeneous point process model (PPM) was then fitted separately for each animal-human interface, as described above in the yellow, orange, and red alert-level zones wherein each interface is included as a single independent variable to examine the fit of, and association between, the interface and the two case-study zoonoses. The PPMs were fit using the spatstat package in R^31^.

Finally, a global network analysis was conducted using the airport adjacency matrix described above to identify highly connected cities that are adjacent to alert-level zones. Network strength, the sum of the weights of a vertex’s edges^32^, and network eigenvector centrality, each vertex’s centrality as proportional to the sum of all of the centralities of its connected vertices^33^, were computed to represent airport network centrality. Network analyses were conducted in R using the igraph package^34^. The airport network is presented in S2 Figure 2, wherein the size of each airport node reflects the degree of centrality (eigenvector centrality). A principal component analysis identified orthogonal measures of network centrality since strength and eigenvector centrality are highly correlated. The principal components (PC) were then used to represent each airport’s centrality in the network. A subset of those cities in the top quartile (75^th^ percentile) of the first principle component of network centrality (90% of network strength and eigenvector centrality variance was explained by the first principle component) was selected to represent cities highly connected to the global airport network (S3 Table 1). Finally, those highly connected cities that were within 50 km of the edge of each alert-level zone were identified as cities at risk of dissemination for high impact spillovers, given their proximity to zones of high-risk interfaces. These high-risk dissemination cities were then mapped along with each of the corresponding alert-level zones. All descriptive analyses were performed in the R environment^35^, except for distance rasters to each of the alert-level zones, which were computed with the proximity raster analysis tool in QGIS^36^. The silhouette images used in figures 1 and 5 were obtained from http://phylopic.org/ and https://publicdomainvectors.org/en/ and are distributed under Creative Commons license.

**Table 1.**
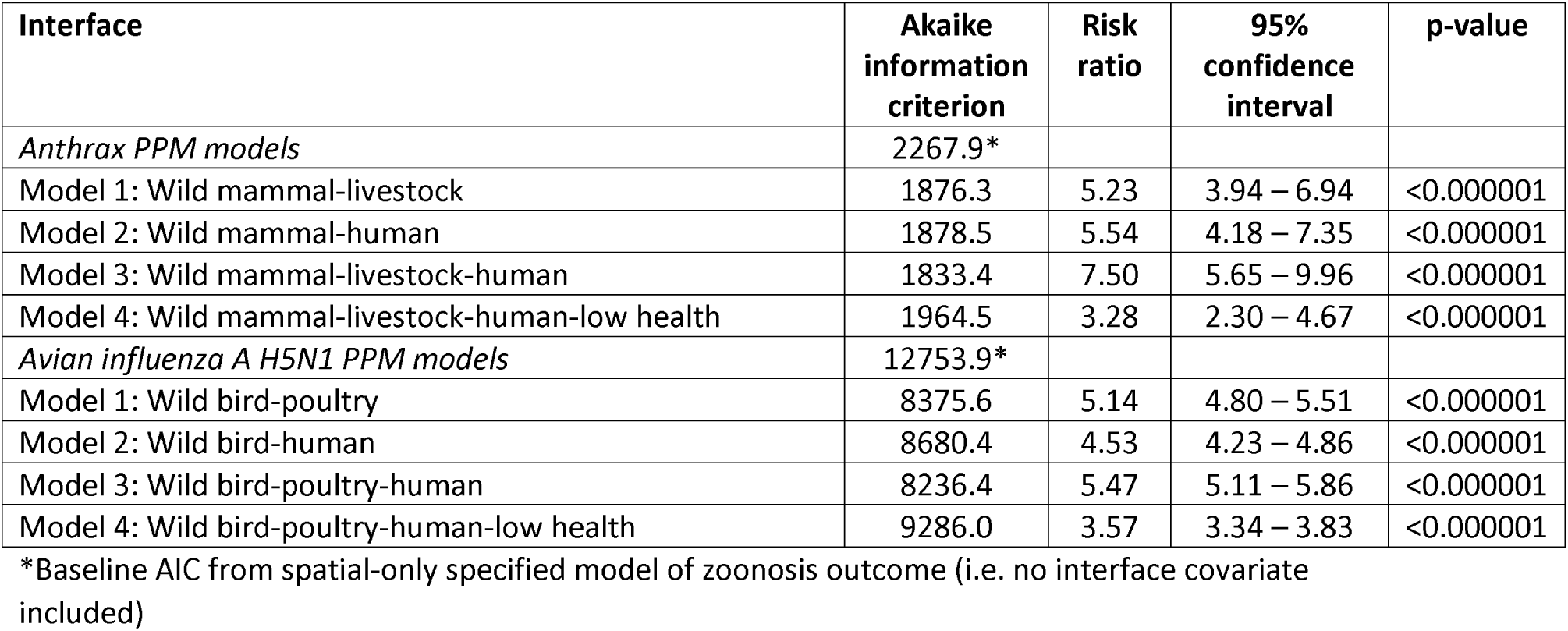
Point process models for each interface under each of the two case studies (anthrax and avian influenza) used to test the validity of these animal-human interface metrics.

**Figure 2.**
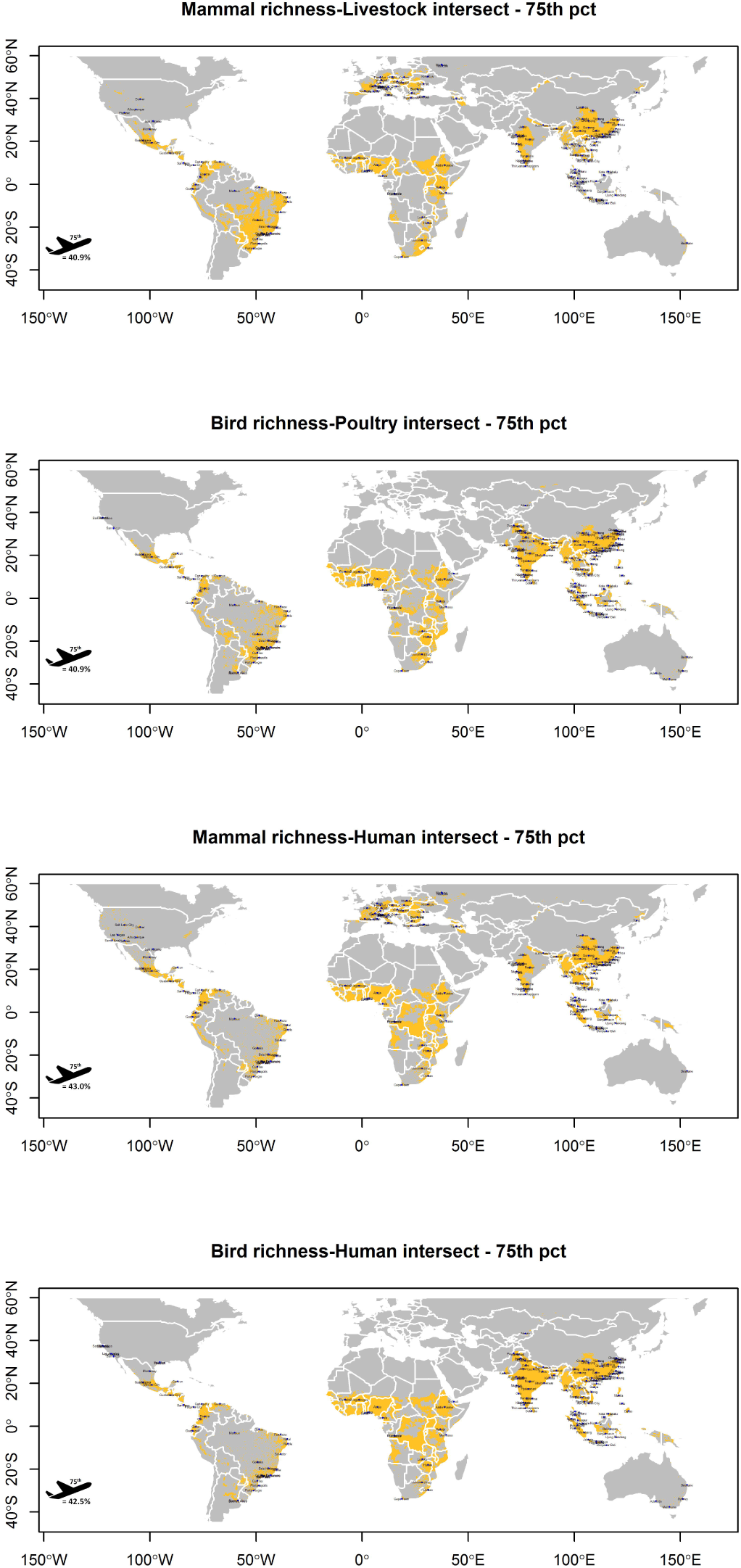
Alert-level yellow zones depict two-way interfaces between mammalian/bird species richness and human population, livestock, and poultry densities, respectively, based on the intersection of the top quartile of each feature’s distribution. Cities of high network centrality (75^th^ percentile) based on global air travel and within 50 km of yellow interfaces are overlaid. The proportion of highly connected transportation hubs (75^th^ percentile of airport network centrality) within 50 km of each interface is presented with the airplane icon. Cities mapped twice have two high centrality airports.

**Figure 3.**
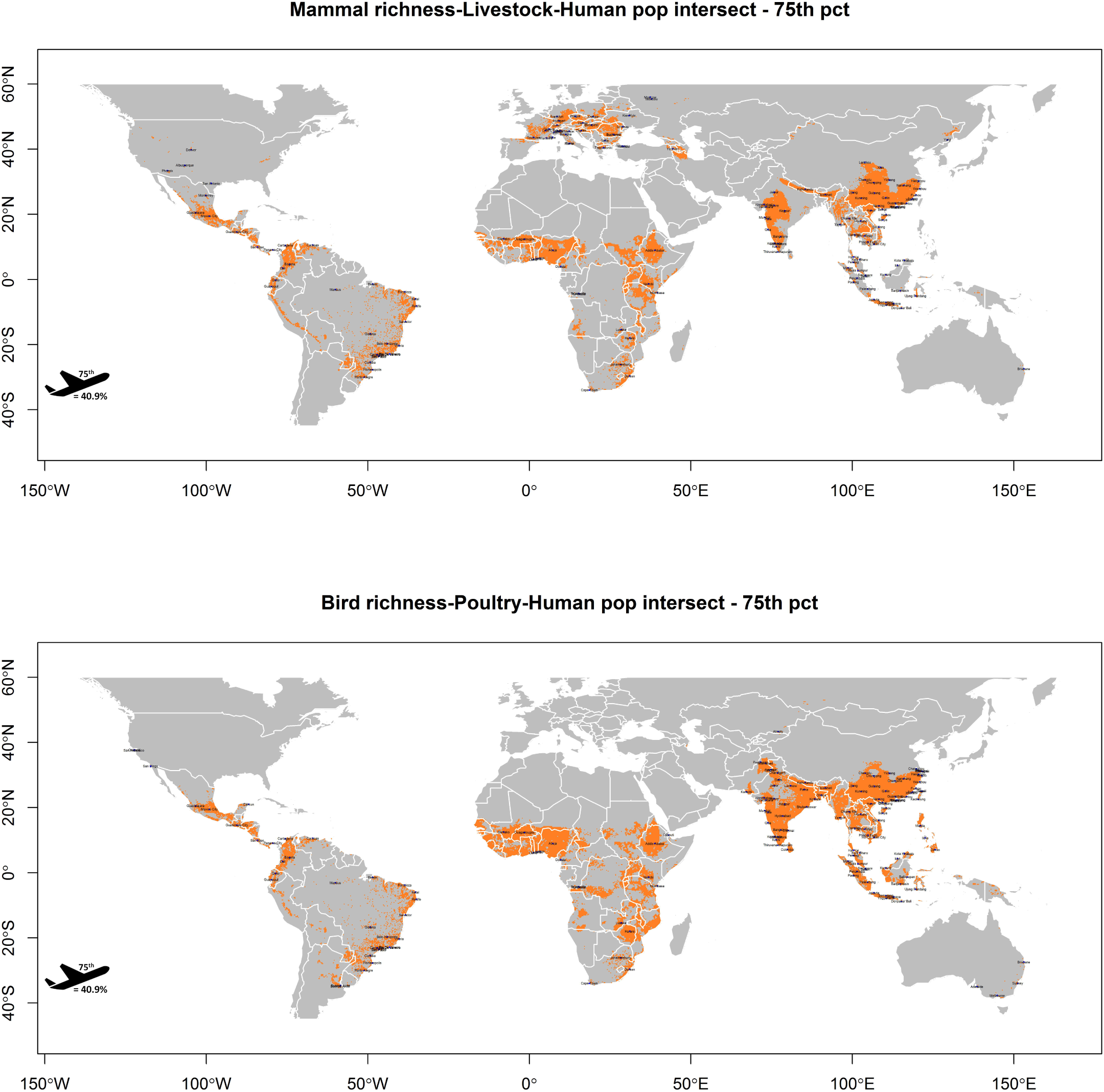
Alert-level orange zones depict three-feature interfaces between mammalian richness, human population density, and livestock/ poultry densities based on the intersection of the top quartile of each feature’s distribution. Cities of high network centrality (75^th^ percentile) based on global air travel and within 50 km of orange interfaces are overlaid. The proportion of highly connected transportation hubs (75^th^ percentile of airport network centrality) within 50 km of each interface is presented with the airplane icon. Cities mapped twice have two high centrality airports.

**Figure 4.**
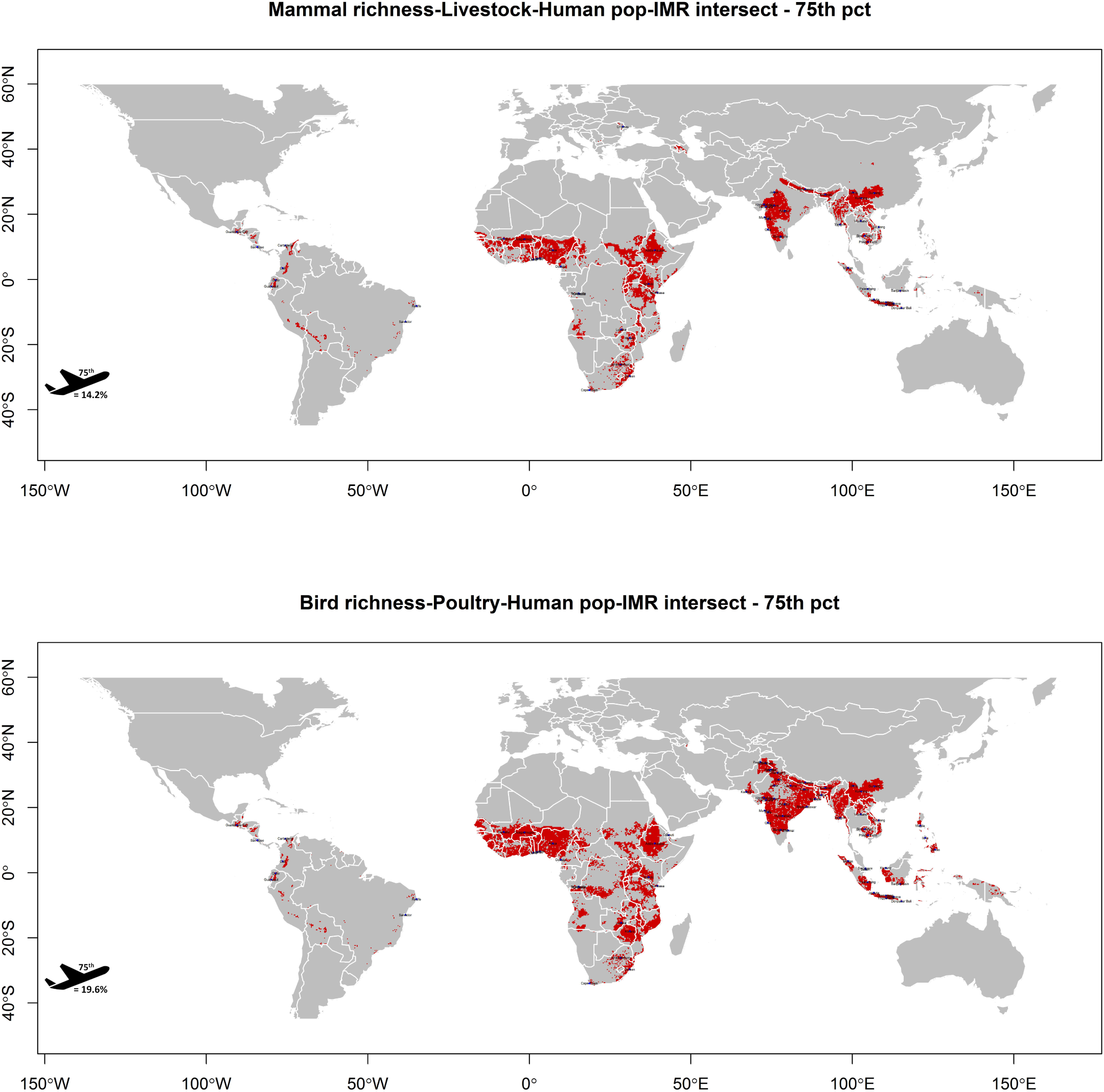
Alert-level red zones depict the same animal-human interfaces as presented in alert-level orange, but extends the intersection of these interfaces with the top quartile of infant mortality. Cities of high network centrality (75^th^ percentile) based on global air travel and within 50 km of red interfaces are overlaid. The proportion of highly connected transportation hubs (75^th^ percentile of airport network centrality) within 50 km of each interface is presented with the airplane icon. Cities mapped twice have two high centrality airports.

**Figure 5.**
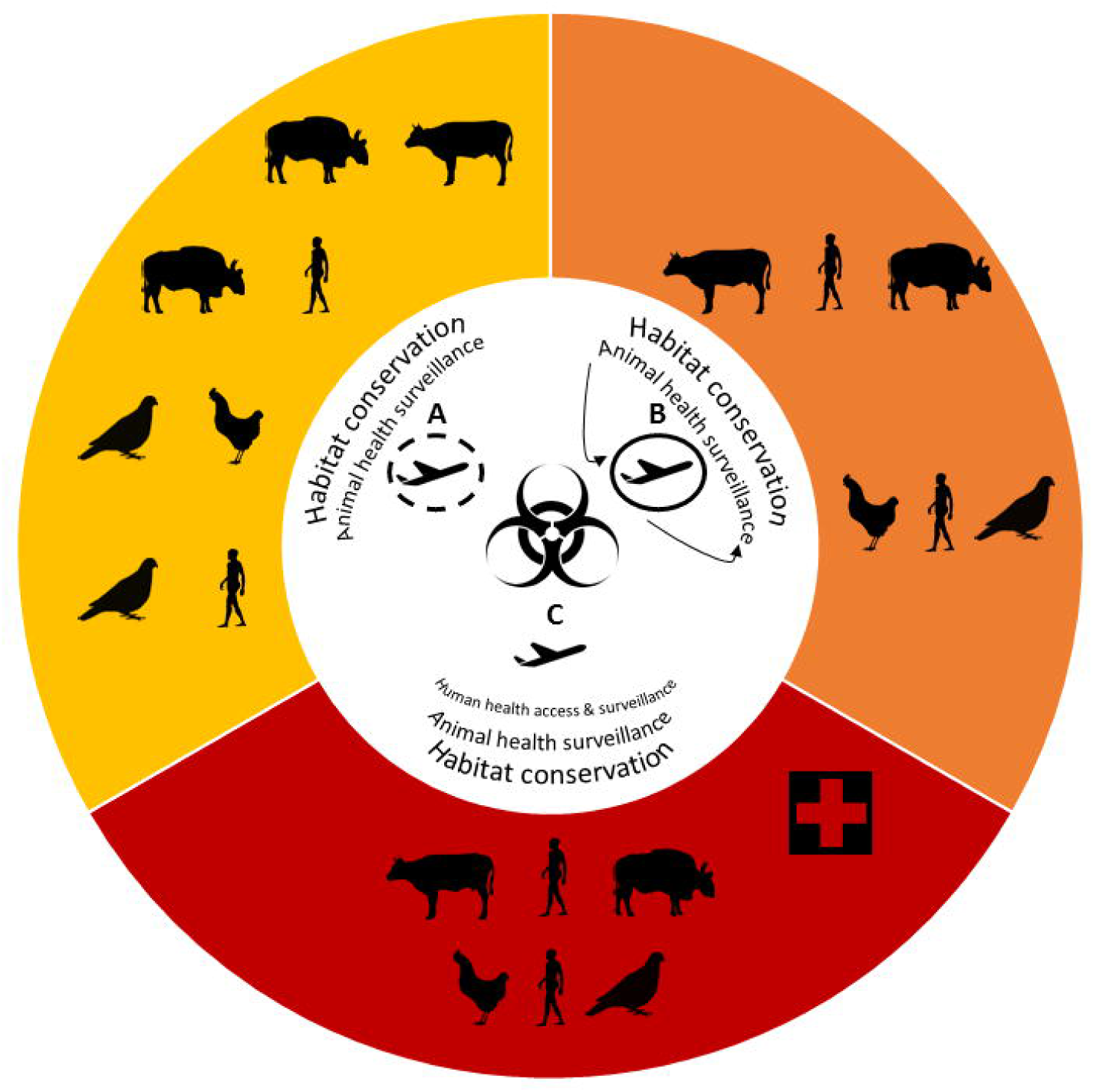
Alert-level zones with barriers to high-impact spillover. All three zones represent landscapes of high anthropogenic pressure intersecting with high biodiversity, and thus all three zones indicate potential for impactful spillover. Interventions are suggested for each zone and represent urgent needs specific to each landscape to block high-impact spillover. Habitat conservation and animal health surveillance are beneficial strategies common to all zones, whereas the red zone additionally requires strengthened health systems and surveillance in humans. The more upstream the strategy, the “closer” it is to the landscape and thus the broader the downstream window of impact (depicted by larger font size). Strategies targeting the transportation hubs alone (A) are the most downstream, i.e. closest to points of global dissemination, and thus provide a narrow window of impact with a permeable barrier from the point of spillover in the landscape to the point of dissemination in the hub. However, strategies targeting transportation hubs that are integrated with upstream forms of surveillance (B) can strengthen the downstream intervention ultimately contributing to an impermeable barrier from the point of spillover to the point of dissemination. The exclusion of transportation hubs from intervention strategies (C) is ill-advised, since their close proximity to landscapes of impactful spillover should compel their involvement as a critical last line of defense against global dissemination.

## Results

The alert-level yellow, orange, and red zones with their proximate (within 50 km) cities of high global connectivity are presented in Figures 2, 3, and 4 (city lists for each alert zone are available at <u>10.6084/m9.figshare.12661184</u>). Each animal-human interface presented in this risk hierarchy was strongly associated with the case study zoonoses and a good fit to the data (Table 1), which supports the validity of the interface metrics. Of note, the alert-level yellow and orange zones were a better fit to both case study zoonoses than were the alert-level red zones, indicating those areas of high animal-human interface with poor health system performance were not capturing all outbreaks and thus manifesting surveillance gaps, as expected. Of those cities that were in the top quartile of network centrality, greater than 40% were within 50 km of both the alert-level yellow and orange zones for each interface represented (Figures 2 and 3). Whereas a reduced but still substantial proportion of these cities were within 50 km of the alert-level red zone for the wild mammal-livestock-human interface and wild bird-poultry-human interface (14.2% and 19.6%, respectively; Figure 4). The interface zones of highest potential spillover impact, and their adjacent cities, were predominantly in sub-Saharan Africa and South and Southeast Asia (Figure 4). All descriptive analyses were repeated at the next quartile (50th percentile) for each interface and city centrality, at which 43% of cities in the top 50th percentile of network centrality were within 50 km of alert-level red zone for each interface, while ≥ 81% of cities were within 50 km of alert-level yellow and orange for each distinct interface (supplemental material: S4 Figure 3, S5 Figure 4, and S6 Figure 5).

## Discussion

This work has defined a hierarchical geography of potential high-impact spillover based on variable animal-human interfaces, human health system capacity and their proximate cities of high global connectivity. This work was a descriptive geographical exercise intended to quantify and map a systematic representation of the global animal-human interface, and subsequently it showed that many of the world’s most connected cities are adjacent to or within areas where wildlife share space with humans and their domesticated animals. Indeed, more than 40% of these cities were within or adjacent to landscapes of extensive animal-human interface, while approximately 14%-20% were located in landscapes of both extensive interface and poor health system performance, thus demonstrating the precarious positioning of many global transportation hubs. As a means toward preventing future high-impact spillovers of pandemic potential, we have highlighted those areas of the world that could most benefit from simultaneous investment in 1) conservation efforts to limit wildlife encounters with humans and their domesticated animals, 2) improved surveillance of animals, and 3) improved human health infrastructure to detect spillovers when they occur and prevent onward spread. Finally, defining a global geography of potential high-impact spillover offers unique value by locating those areas and cities that could most benefit from the development of composite, or at least coordinated, landscape and airport biosurveillance systems at local and national levels.

In this study we identify areas where the sharing of space between wildlife and humans is high, health system performance is low, and critical interfaces are located adjacent to cities with high global connectivity. We thus provide practitioners in human and animal health with a geographical framework that applies a synthesis of epidemiological, ecological and health system research outputs to aid evidence-based public health decision-making. We emphasize that this work is not presented as a prediction of global hotspots of spillover. Several well-conducted studies have been undertaken that provide useful results in this domain^5,37,38^. Moreover, previous efforts have clearly demonstrated the importance of increasing human and domesticated animal encounters with wildlife as a key driver of spillover^39^, as well as the potential modulation of wildlife-human encounters and subsequent spillover by climate change^40^. While these previous modelling studies have identified the importance of human pressure on wildlife for spillover, none have described the global distribution of animal-human interface as a data-driven construct of the degree of shared space between wildlife, domesticated animals, and humans. Nor have they considered the location of proximate transportation hubs, which are positioned to amplify impactful spillovers through rapid regional or global dissemination and thus transitioning impactful spillovers to high-impact spillovers with regional epidemic or global pandemic consequence. In other words, conduits of high-impact spillover have not been previously systematically mapped, nor have they been described using a practical hierarchy designating the degree of animal-human interface. The practical approach presented here will allow for a more targeted development of One Health surveillance and prevention programs, and provides considerable scope as a strategic tool for preventing future pandemics. For example, a preventive strategy can be designed to create barriers between the high-hazard landscapes and their adjacent points of global dissemination. Moreover, the closer an intervention is to the landscape generating the hazard (i.e. spillover) the broader would be its expected downstream prevention effect, whereas an intervention implemented farthest from initial spillover (e.g. at transportation hubs) is more permeable and would expectedly provide narrower effect. However, when integrated with upstream surveillance, enhanced monitoring at transportation hubs can contribute to robust, multi-tiered systems by providing a data-informed, critical last line of defense (Figure 5).

Some limitations with this work regarding proxy measures require further discussion. First, the animal-human interfaces described here are based on the distributions of the species involved (wild mammals and birds, domesticated animals, and humans) rather than on direct observations of the species’ interactions in the landscape. Therefore the metrics used to represent these interfaces are proxies for interspecific interaction. Nevertheless, the granularity of the species’ distributions was relatively fine scale (10 km × 10 km) and, given the high percentile (75^th^) of species’ distributions used to construct the primary risk zones, it is reasonable to assume that humans and animals do indeed share the spaces, either in whole or in part or directly or indirectly, within the interfaces we have demarcated. Second, the infant mortality rate was used as a proxy for health system performance. We recognise that health systems are complex coalescences of economic and social infrastructure, medical capacity and training, public health capacity and training, governmental organization, and the general socioeconomic status of human populations. Moreover, it would not be possible to measure these individual components with the granularity required to synthesise a geographically meaningful new construct of health systems. However, the infant mortality rate has long been recognised as both a reliably measured outcome of health systems and a robust metric for comparing the performance of health systems between countries^19–21^. Finally, the validation case studies should not be over-interpreted with respect to anthrax or HPAI H5N1 or any other zoonosis. As described above, the current work is not intended as a predictive modelling study. The models used here were not trained with one set of outbreaks to identify some optimal suite of predictors, and then, once identified, tested and evaluated against an independent set of outbreaks. Rather, under the current framework specific hazard metrics for animal-human interfaces were created by quantifying the extent to which humans share space with wildlife. The point process models were then used to determine how these metrics, data-driven and standradised but conceptualised a priori, actually fit against real-world outbreaks of important known zoonoses. It is also important to recognise that outbreak reporting as captured by the EMPRES-i system may not be geographically homogeneous, which is in fact reflected in the finding that red zone interfaces were a poorer fit to the case study outbreaks indicating that these areas may be more prone to missing cases, as expected.

In conclusion, this investigation has shown that there are substantial animal-human interfaces in areas of poor health system performance, highlighting those areas of potential impactful spillover where health infrastructure may be insufficient to identify spillover cases early and block onward human-to-human transmission should this emerge. This work further showed that there are many cities with a high degree of global connectivity that are proximate to the areas at risk of impactful spillover, and thus has identified the geographic distribution of conduits of potential high-impact spillovers. One Health practitioners can now apply this to the essential work of bolstering disease surveillance at the animal-human interface and the adjacent hubs of global air travel to protect the world from future pandemics.

## Data Availability

Data are available at the Figshare link provided.

https://figshare.com/articles/High-impact_spillover_landscapes_and_cities/12661184

## Acknowledgements

The current COVID-19 pandemic constitutes one of greatest challenges to our collective human experience. The authors would like to acknowledge and express their gratitude for all of the heroic efforts of the health care workers, public health workers, and essential services workers, who have met this incredible challenge with the resolve, strength, and compassion that defines the best of humanity. Without their efforts, we would not have been afforded the safe space in which to conduct this work. Their contribution to the future of scientific endeavor, and indeed all human endeavor, cannot be overstated and must never be forgotten.

## Author contributions

MW contributed to conceptualisation, design, data analysis, and draft writing; SS contributed to conceptualisation, design, and draft writing; SH contributed to conceptualisation, design, and draft writing; SM contributed to conceptualisation, design, and draft writing.

## Materials & correspondence

Please address correspondence and material requests to Michael Walsh

